# A new transmission route for the propagation of the SARS-CoV-2 coronavirus

**DOI:** 10.1101/2020.02.14.20022939

**Authors:** Antoine Danchin, Patrick Tuen Wai Ng, Gabriel Turinici

## Abstract

**Background:** A novel coronavirus (SARS-CoV-2) spread from the capital of the Hubei province in China to the rest of the country, then to most of the world. To anticipate future trends in the development of the epidemic, we explore here, based on public records of infected persons how variation in the virus tropism could end up in different patterns, warranting specific way to handle the epidemic.

**Methods:** We use a compartmental model to describe the evolution of an individual through several possible states: susceptible, infected, alternative infection, detected and removed. We fit the parameters of the model to the existing data taking into account significant quarantine changes where necessary.

**Results:** The model indicates that Wuhan quarantine measures were effective but that alternative virus forms and a second propagation route are compatible with available data. For Singapore and Shenzhen region the secondary route does not seem to be active yet and the epidemic size limited.

**Conclusions:** The alternative infection tropism (the gut tropism) and a secondary propagation route are validated hypotheses using a model fitted by the available data. Corresponding prevention measures that take into account both routes should be implemented to the benefit of epidemic control.

## Introduction

Late in 2019 a novel coronavirus (named SARS-Cov-2 later on) was detected in mainland China originating from the Wuhan province. The importance of the disease took some time to be acknowledged^1,2^, resulting in a fairly significant number of infected persons who subsequently spread the disease throughout China^3^ (all provinces had at least one case on January 30th), then world-wide. This makes it essential to explore the way the epidemic may spread in the near future. In the present work we proposed several scenarios to this aim. While it is obviously hazardous to advance models of epidemics before their course has been completely unfolded, we think that it is helpful to evaluate methods meant to understand their ongoing course. In particular by following how the epidemic developed at places other than its original site of onset, we would be able to detect any unexpected development course that might be used by health authorities to react very early on. This would be extremely useful to detect patterns created by socio-political measures meant to contain the disease or mutants of the virus which would result into higher contagion or virulence, thus prompting a rapid response.

It has been established that coronaviruses are versatile in their preferred site of infection. These viruses have the option to pass from a “gut tropism” version to a “lung tropism” instance, as was observed for other coronavirus outbreaks).^4–7^ >Depending on the infected person or virus spread dynamics, coronavirus effects can be more or less severe: the virus can either preserve its lung tropism with adverse impact or act as a “gut tropism” version and be relatively harmless. First reports established that the novel 2019 instance also induces gut symptoms^8^. Its genetic build up and evolution is still subject to intense research^9^. This multifaceted behaviour may result in unexpected local courses of the epidemic as suggested in a scenario of a “double epidemic”, as proposed for the SARS 2003 worldwide epidemic^10^, except that in the present situation we would be witnessing the effect of a single epidemic with modulated effects and propagation depending on the affected patients and random virus mutations.

### A secondary propagation route

Due to the versatility of the virus, in addition to interfering with the immune response, contagion may involve a variety of causes, such as environments contaminated with virus carrier secretions, dirty water effluents, besides the expected direct contamination via aerosols. This makes that the oro-fecal route should be considered as an important complement of contact with the virus (recent information sustains this hypothesis^11^).This was observed at the Amoy Gardens cluster of cases for SARS^12,13^. This observation prompted us to include in our model, besides the major respiratory route, a secondary propagation route, which is not a direct human-to-human propagation but rather some indirect vector (or environment element) to/from human. A second important consequence of assuming the presence of an indirect route is that the selection pressure on virus mutants will differ considerably between lung tropism and gut tropism.

Finally, there may be a difference in incubation time, the “gut tropism” version of the virus would possibly cause less fever^14^. This hypothesis is consistent with some propagation patterns, e.g. the first case in Macau, which was not detected at the border, implying that that the affected person did not have fever.

We should note that this makes the disease considerably more dangerous in terms of propagation because carriers are, at least for some time “invisible” and display a risky behavior^15,16^.

This explains our choice of compartmental model (see below). Nevertheless, it appears that some people were infected and are already discharged from hospitals, which leads to conclude that the disease is, at this time, less dangerous than SARS.

We propose several scenarios compatible with past scenarios of coronavirus propagation in parallel with our propagation model. Besides providing an estimation of the impact of the epidemic, this work also suggests some countermeasures to slow down propagation. Compared with SARS, the death burden could become much higher (on a par with what is observed with flu) because of lack of proper containment.

## Methods

The mathematical model of epidemic propagation is the following (see Figure 1):

**Figure 1.**
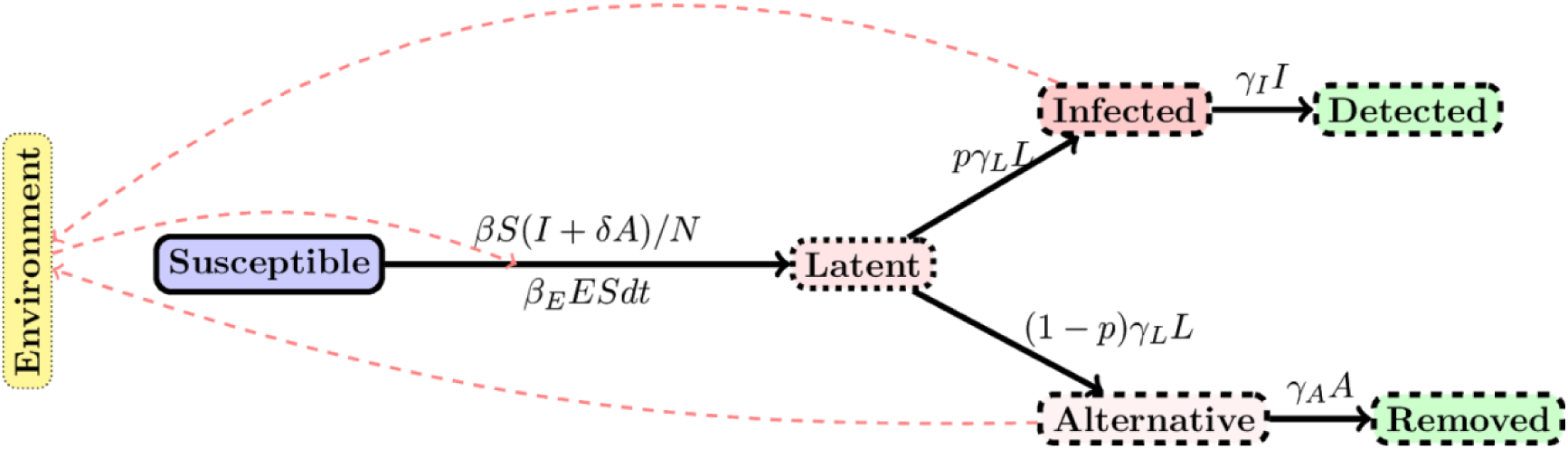
Schematic view of the SLIADRE model used in the simulations. Each compartment is indicated with a rectangular box and the flow of individuals or infection vectors with solid arrows. Dashed arrows indicate which compartment influences which flow.

**Figure 2.**
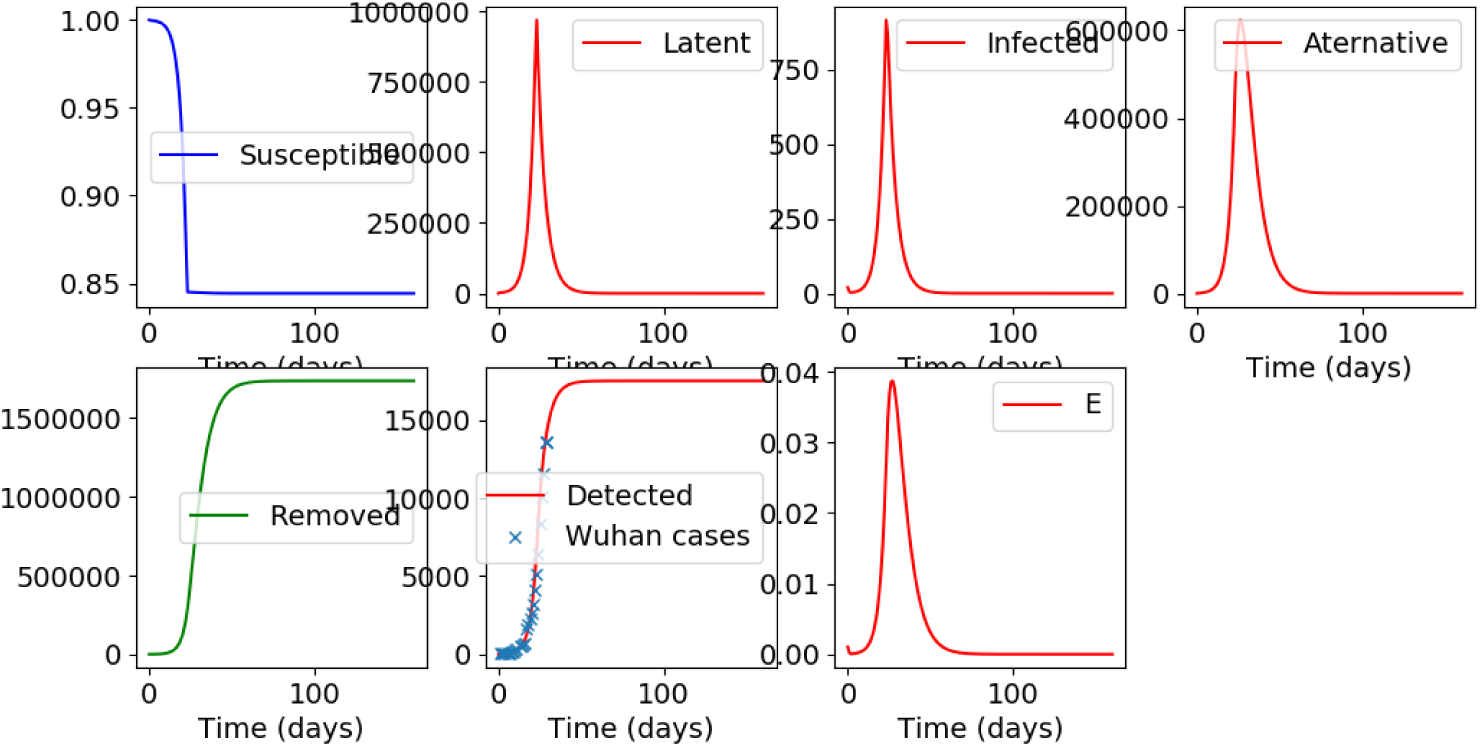
Results of the model for Wuhan city epidemic.

**Figure 3.**
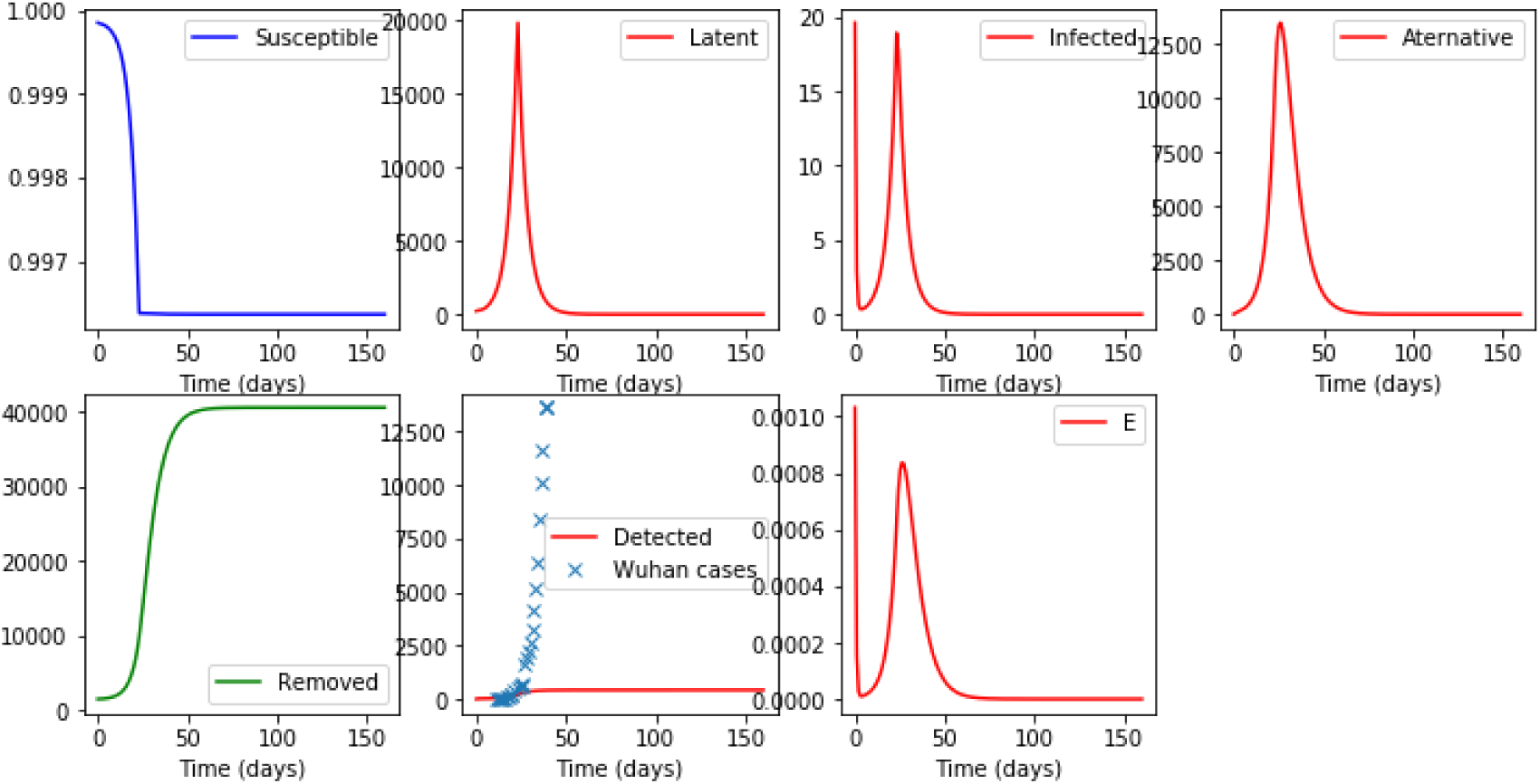
Results of the simulations when the environmental propagation route is switched off. The classical propagation route alone induces a much smaller epidemic size than actual data indicates.

**Figure 4.**
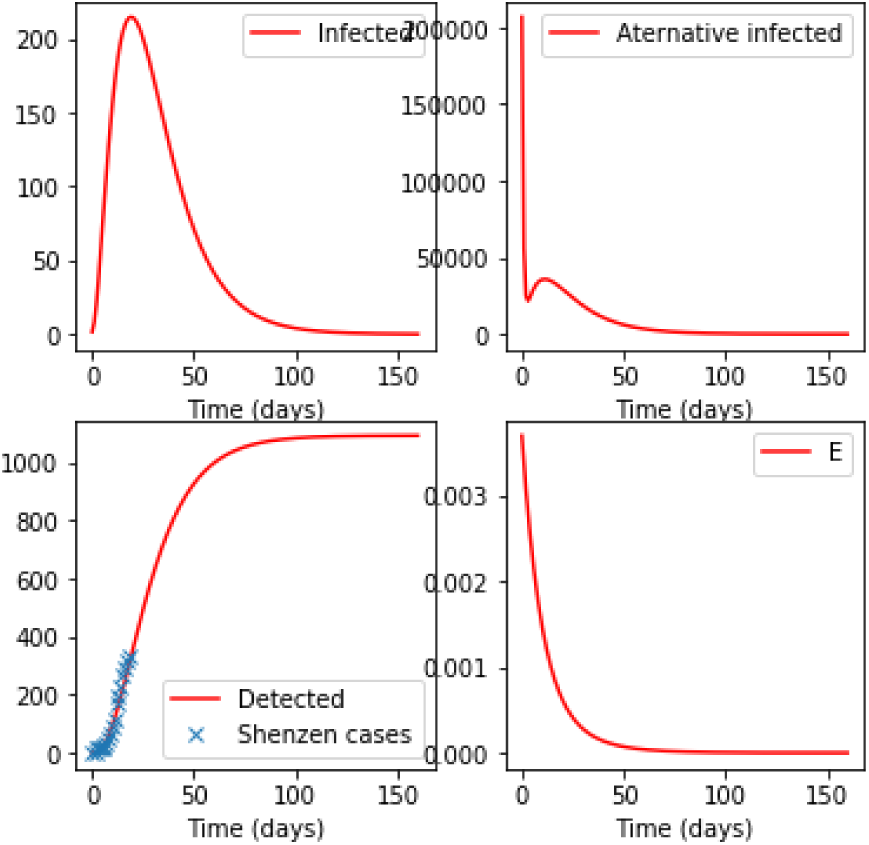
Results of the SLIADRE model for Shenzen.

- we use a compartmental model^17,18^ to describe the evolution of an individual through several possible conditions such as susceptible (compartment label “S”), latent, i.e. infected but not yet infective (label “L”), infective in the lung tropism and highly symptomatic (label “I”), low symptomatic, infective in the gut tropism (“A”); the “I” infected then go to “Detected” (D) stage while the “A” infected go to “Removed /recovered” stage (label “R”). This is an adaptation of existing models and especially the SLIAR model^19–23^.
- We add to the model the possibility for an environmental, local, propagation route (that accounts for soiled water, etc.). This option is represented by the “E” label. With some probability, the contact between infected environmental elements and a susceptible individual gives rise to new infections.

Here N is the total population; the parameters β, β_EI_, β_ES,_ δ, γ_L,_ γ_I_, γ_A_, γ_E,_ and p, of the model are chosen in order to be compatible with past works and present insights^24^. However since the public and health authorities responses are not known and not anticipated, some uncertainty remains in the parameters which leads us to consider several choices of the parameter values giving rise to several scenarios of epidemic propagation.

The mathematical formulation of the model is:

dS/dt = -β*S*(I+ δA)/N - β_ES_ *E*S

dL/dt = β*S*(I+ δA)/N + β_ES_ *E*S – γ_L_*L

dI/dt = p*γ_L_*L - γ_I_*I dA/dt = (1-p)*γ_L_*L - γ_A_*A dD/dt = γ_I_*I

dR/dt = γ_A_*A

dE/dt = β_EI_*(I+ δA)/N-γ_E_*E

## Results

We start with a simulation of the Wuhan city epidemic. We take into account the quarantine starting January 23rd and impose a reduction (given by a multiplicative factor that is fitted numerically) of the transmission parameters β and β_ES_ following that date. This attenuation is found, as is the case for all other parameters, in the course of a search procedure that imposes a match of the “Detected” patients of the model and the number of reported cases available from communications of the WHO or Chinese authorities. The results are displayed below for the optimal parameters given in the supplementary material. The epidemic size is around 32,000. The key observation here is the presence and importance of the alternative form “A” and the alternative propagation route ES (see also below).

The effect and importance of quarantine measures: we tested what happens if we discard the attenuation factors found by the fit procedure. We obtained a total epidemic size of 112,539, which is a sharp increase with respect to the baseline scenario.

### The importance of the alternative form and the alternative propagation route

In this scenario, we neglected the alternative propagation route by setting the β_ES_ parameter to zero in the previous run. We obtained the figure below, which shows that absence of the ES propagation route cannot explain the way the epidemic is unfolding (total epidemic size is down to 415 from previous figure of 17,500, closer to observations).

Similar considerations hold when switching down the Alternative form of virus presentation (results not shown here). We continue with a simulation of the propagation of the epidemic in Shenzhen: the epidemic seems to be relatively under control with a final epidemic size of around 1,000.

The next simulation explores the case of Singapore: the results (not shown here) are similar to those of Shenzen, there the epidemic does not appear to be self-sustained; the model predicts an epidemic size of 130.

## Discussion

A word of caution concerning the present view: any model is but an imperfect view of reality and this model is no exception. In addition, the quality of the results given by the model depends directly on the quality of the data input, and it seems likely that some of the reported figures are subject to large error bars; in particular all simulations of this model were done before February 12^th^ when the accounting procedure for of number of cases was changed in China. Therefore, one should take the exact figures obtained with our simulations with a grain of salt. Yet, the simulation resulting from the model does show some important qualitative features that are now discussed.

Specifically, the hypothesis that there exists a secondary route of transmission is meaningful and that two major types of virus effects are present is supported by the simulation data. This has the interesting consequence that it appears that, for the city of Wuhan, the quarantine seems to be effective—of course notwithstanding the need for further quarantine efforts to ensure that it remains under control—. However, since a reservoir is probably present, and the oro-fecal route may be an important propagation factor, the prevention of this transmission element is vital to avoid any resurgence. **The first recommendation seems to be the enforcement of strict post-epidemic measures at the possible reservoir sites**.

In Singapore (and partially in Shenzhen as well) even if the current data indicates that the number of cases is likely to increase, is does not indicate that a secondary propagation route is already effective. However, efforts are to be made to ensure that this remains true in the future and the control of the secondary route, which makes the difference between a large scale epidemic and a controlled outbreak, remains an important target for public health measures.

## Conclusions

Elaborating on the behavior of previous coronavirus outbreaks, we worked out the hypothesis that an alternative infection tropism (the gut tropism) linked to a secondary propagation route (through environment) is affecting the development of the present 2019-nCoV epidemic. Our epidemic propagation model, when fit to existing data, indicated that, among all regions analysed (Wuhan city in the Huabei region in mainland China, Singapore, Shenzhen region), the propagation of the disease in the city of Wuhan underwent an original course. It appeared to be substantially facilitated by a secondary propagation route, thus substantiating the beneficial effect of an effective quarantine. The main message of our exploration is that relevant prevention measures that take into account both propagation routes should be implemented to contain the extension of the epidemic to further sites, especially when novel sites are uncovered.

## Data Availability

no personal of private data were used in this work.

